# Relevance of brain MRI in patients with uveitis: retrospective cohort on 402 patients

**DOI:** 10.1101/2020.03.22.20021113

**Authors:** N. Chirpaz, S. Kerever, A. Gavoille, L. Kodjikian, R. Bernier, M. Gerfaud-Valentin, P. Denis, T. Mathis, Y. Jamilloux, P. Sève

**Affiliations:** Department of Internal Medicine, Hôpital de la Croix Rousse, Hospices Civils de Lyon, F-69004, Lyon, France; Department of Ophthalmology, Croix-Rousse Teaching Hospital, Hospices Civils de Lyon, Lyon, France; Department of Anesthesiology and Critical Care, Lariboisière University Hospital, AP-HP, University of Paris France; UMR-CNRS 5510 Matéis, Villeurbanne, Université Claude Bernard Lyon 1, University of Lyon, Lyon, France; Hospices Civils de Lyon, Pôle IMER, Lyon, F-69003, France; Univ. Lyon, University Claude Bernard Lyon 1, HESPER EA 7425, F-69008 Lyon, France

**Author notes:** contributed equally to this work (S. Kerever, A. Gavoille). **Corresponding author:** Prof. Pascal Sève, Department of Internal Medicine, Hôpital de la Croix-Rousse, 103 grande rue de la Croix-Rousse, F-69004, Lyon, France. **Synopsis** Although brain magnetic resonance imaging (bMRI) is frequently performed as part of the etiological assessment of uveitis, very few prior studies have evaluated the relevance of this exam.

**Keywords:** brain magnetic resonance imaging, uveitis, multiple sclerosis, oculocerebral lymphoma, retinal vasculitis

## Abstract

**Aim:** To assess the diagnostic value of brain magnetic resonance imaging (bMRI) for the etiological diagnosis of uveitis and to establish predictive factors associated with its advantageous use.

**Methods:** Retrospective study on all patients with *de novo* uveitis who were referred to our tertiary hospital and who underwent a bMRI between 2003 and 2018. The bMRI was considered useful if it served to confirm a diagnosis or correct a misdiagnosis. We also collected characteristics of uveitis and associated ophthalmological and neurological clinical signs.

**Results:** Brain MRI was contributive in 19 out of 402 cases (5%): 10 multiple sclerosis, 5 radiologically isolated syndromes, and 4 oculocerebral lymphomas.

A total of 34 (8%) had neurological signs and 13 (38%) of those patients had a contributive bMRI. Meanwhile, in the absence of neurological signs, 1% of bMRIs were contributive, and none of them resulted in specific treatment.

Among patients with a contributive bMRI, 68% had neurological signs. Univariate analysis established that neurological signs (p<0.001), granulomatous uveitis (p=0.003), retinal vasculitis (p=0.002), and intermediate uveitis (p<0.001) were all significantly associated with a contributive bMRI. Multivariate analysis confirms the significant association of neurological signs (p<0.001) and intermediate uveitis (p=0.01).

Patients with oculocerebral lymphoma were significantly older (p<0.001) and all were above 40 years of age.

**Conclusion:** Brain MRI appears to be a relevant and contributive exam, but it should be performed in cases of intermediate/posterior uveitis or panuveitis accompanied by neurological signs, retinal vasculitis, or in patients older than 40, to rule out an oculocerebral lymphoma.

## Introduction

Uveitis is an intraocular inflammation that affects both the uveal tract and adjacent structures such as the vitreous and the retina. It is the third cause of blindness worldwide and currently accounts for approximately 10% of preventable vision loss in the United States and up to 15% worldwide [1,2]. The underlying causes are multiple and mainly represented by inflammatory diseases, followed by pure ophthalmological entities and infectious diseases [3,4].

Although the etiological diagnosis of uveitis is important for prognosis and therapeutics, the etiology remains uncertain or unknown in 28 to 45% of cases [1,5–7], depending on the anatomic type of uveitis. Due to the multiple underlying causes and its heterogeneous presentation, etiological diagnosis is a challenge for patient care and health economics [1,8]. So far, the utility of diagnostic tests has mainly been evaluated in retrospective studies and often in isolated or separate manner [9,10]. One prospective study named ULISSE (Uveitis: clinical and medicoeconomic evaluation of a standardized strategy for etiological diagnosis) has shown that a standardized strategy for the etiological diagnosis of uveitis was neither inferior nor superior to an open strategy [11]. Subsequent analysis has shown that only a few diagnostic tests were useful for the etiological assessment of uveitis [12,13]. These were often cheap, simple, usually guided by clinical findings, and resulted in an etiological diagnosis for many patients.

The benefits of invasive as well as more complex investigations such as brain magnetic resonance imaging (bMRI) could not be determined since these tests were rarely performed. Only a few studies have assessed the diagnostic value of bMRI in uveitis patients [9,14] and to date, patients who may benefit from this exam have not been clearly defined.

In this study, our aim was to assess the usefulness of MRI in the etiological diagnosing of uveitis in a large cohort of patients and to establish predictive factors associated with its advantageous use.

## PATIENTS AND METHODS

### Patients

This study was a retrospective analysis of the records of 1,214 patients with “uveitis” referred by the Department of Ophthalmology (Lyon University Hospital, Lyon, France) or referred by non-hospital ophthalmologists to the department of Internal Medicine (Lyon University Hospital, Lyon, France) between January 2003 and July 2018. The uveitis diagnosis was always confirmed by an ophthalmological examination. Cases of uveitis occurring in the course of previously-diagnosed diseases were excluded. This study was approved by the ethics committee of the French Society of Ophtalmology (IRB 00008855 Société Française d’Ophtalmologie IRB#1).

### Diagnostic work-up and definitions

All the patients underwent a standard screening protocol for uveitis, which included a C-reactive protein test, a complete blood cell count (CBC), a serological test for syphilis, and a chest X-ray [3,11]. Human leucocyte antigen typing (HLA)-B27 was performed in patients with acute anterior uveitis. In patients with chronic anterior uveitis or granulomatous uveitis, a measurement of blood angiotensin-converting enzyme (ACE), a tuberculin intradermal reaction and/or a QuantiFERON TB Gold Plus test, and a chest computerized tomography (CT) scan were performed.

Diagnostic screening for sarcoidosis included conjunctival or skin biopsies if there were clinically suggestive features. Some patients underwent a minor salivary gland biopsy (MSGB), a transbronchial lung biopsy, a bronchoalveolar lavage (BAL) or a PET-scan imaging. This work-up was completed in some patients by an anterior chamber paracentesis (with a polymerase chain reaction for Herpes virus, *Toxoplasma* or RNA16S, and sometimes an interleukin-10 measurement), and/or a vitreous biopsy. bMRI and lumbar puncture were mostly performed in the presence of intermediate or posterior uveitis, or panuveitis. Brain MRI was considered contributive when there were useful findings for etiological diagnosis, and non-contributive when it was normal or in the presence of non-specific abnormalities unrelated to the etiology.

The Standardization of Uveitis Nomenclature was used throughout this study for the anatomical classification of uveitis [4,15].

Briefly, the diagnostic criteria used were the following:

- the international study group for Behcet’s disease criteria [16];
- the Levinson’s criteria [17] or the global diagnostic criteria for birdshot retinochoroidopathy. All birdshot retinochoroidopathy were HLA-A29 positive;
- the 2010 revised McDonald’s criteria for multiple sclerosis (MS) [18] and Okuda’s criteria for radiologically isolated syndromes (RIS), defined as asymptomatic subjects with bMRI abnormalities suggestive of MS [19]. All MS or RIS diagnoses were made in conjunction with a neurologist.
- the international criteria for the diagnosis of sarcoidosis [20] (with Zajicek’s classification for neurosarcoidosis) [21]. In the absence of histological proof, we used Abad’s modified criteria [22]. Patients were presumed to have sarcoid uveitis if they had at least 2 of the following 4 criteria: typical changes on chest X-ray or CT scan, a predominantly CD4 lymphocytosis on BAL fluid analysis, an elevated ACE or an 18-fluorodeoxyglucose (18-FDG) uptake on scintigraphy. They were judged to have undetermined sarcoid uveitis when only one criterion was met.
- the Assessment of SpondyloArthritis international Society (ASAS) criteria for spondyloarthritis [23]. HLA-B27 positive patients with acute non-granulomatous anterior uveitis without clinical and/or radiological features of spondyloarthritis were diagnosed as having HLA-B27-related anterior uveitis;
- the revised diagnostic criteria for Vogt-Koyanagi-Harada (VKH) disease [24];
- Gupta’s criteria for the diagnosis of intraocular tuberculosis [25].

Whenever the intraocular inflammation could not be assigned to a specific diagnosis, the uveitis was considered to be idiopathic.

### Data collection

We collected patients’ demographic data, follow-up times, and the following ophthalmological characteristics at diagnosis: anatomical type of uveitis (anterior, intermediate, posterior, pan uveitis) [11] and anterior chamber examination (tonometry, slit lamp, biomicroscopy to assess whether the uveitis was granulomatous or not). We also reported whether the uveitis was acute or chronic, and uni- or bilateral.

### Statistical analysis

The data are described as their frequencies and percentages for the categorical variables, and as their medians (25th–75th percentile range) for the quantitative variables. Categorical variables were compared using the Chi-square test or the Fisher exact test, as appropriate, and quantitative variables using Wilcoxon’s ranked-sum test. After assessment of possible colinearity between variables by using variance inflation factor, the logistic regression model was built by using backward stepwise selection method with contributive bMRI as the dependent variable. Independent and clinically relevant predictors selected from univariate analysis (p < 0.2) were entered into the model by using rule of 10 events per variable.

All tests were two-sided and statistical significance was set at the p□=□0.05 level. Analyses were performed using R open source software 3.4.4 (available online at http://www.R-project.org).

## RESULTS

### Patients characteristics

After excluding 312 patients because of insufficient/missing data, 903 patients were included, of whom 402 patients (44.5%) underwent a bMRI. Baseline characteristics of these 402 patients are summarized in Table 1. Overall, the mean age at diagnosis was 49 years (range, 31-62) and 230 were women (57%). Sixty-four patients (16%) were non-Caucasians, including patients from North Africa (12%), Sub-Saharan Africa (2%) and Asia (1%).

**Table 1:**
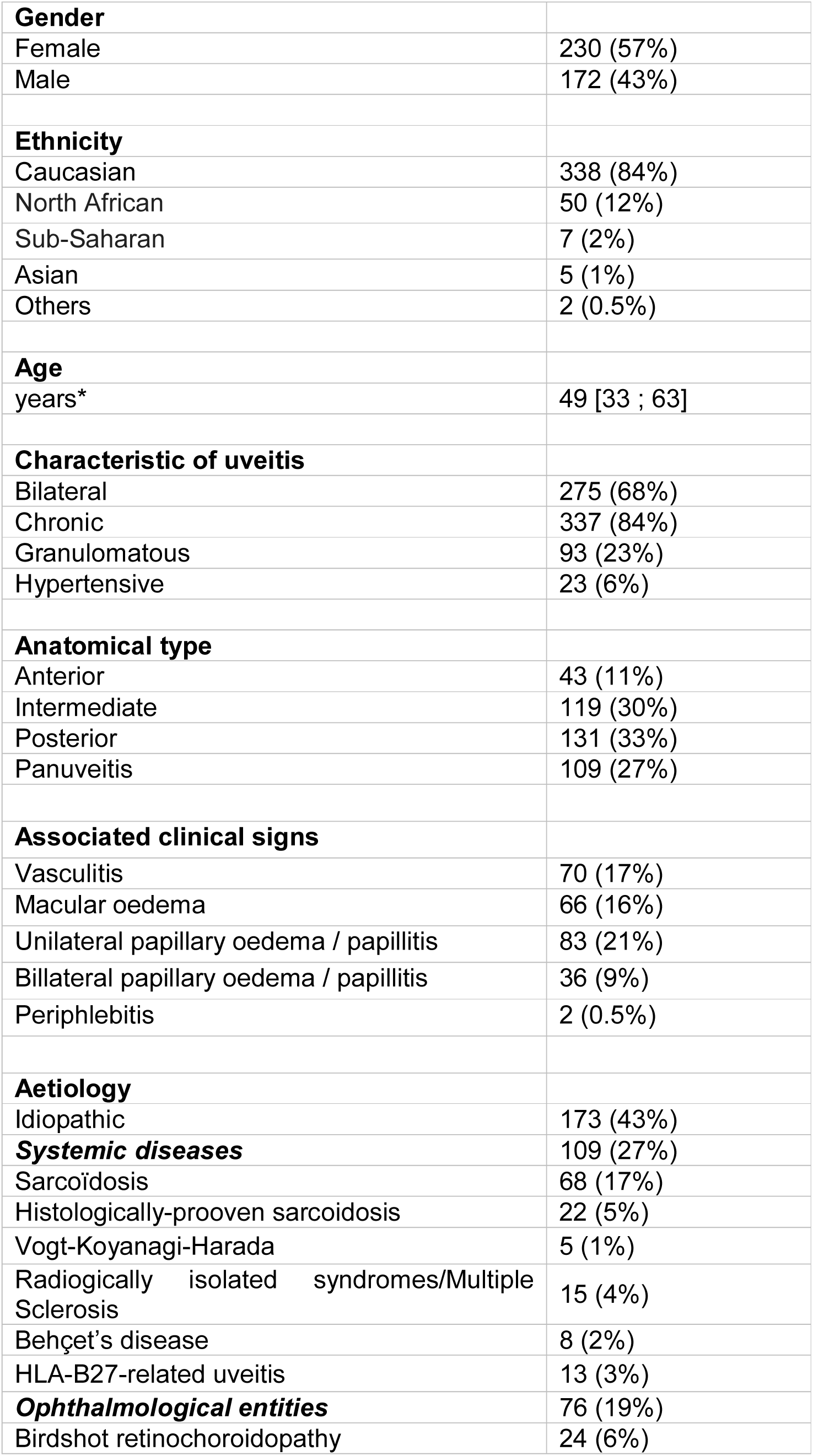
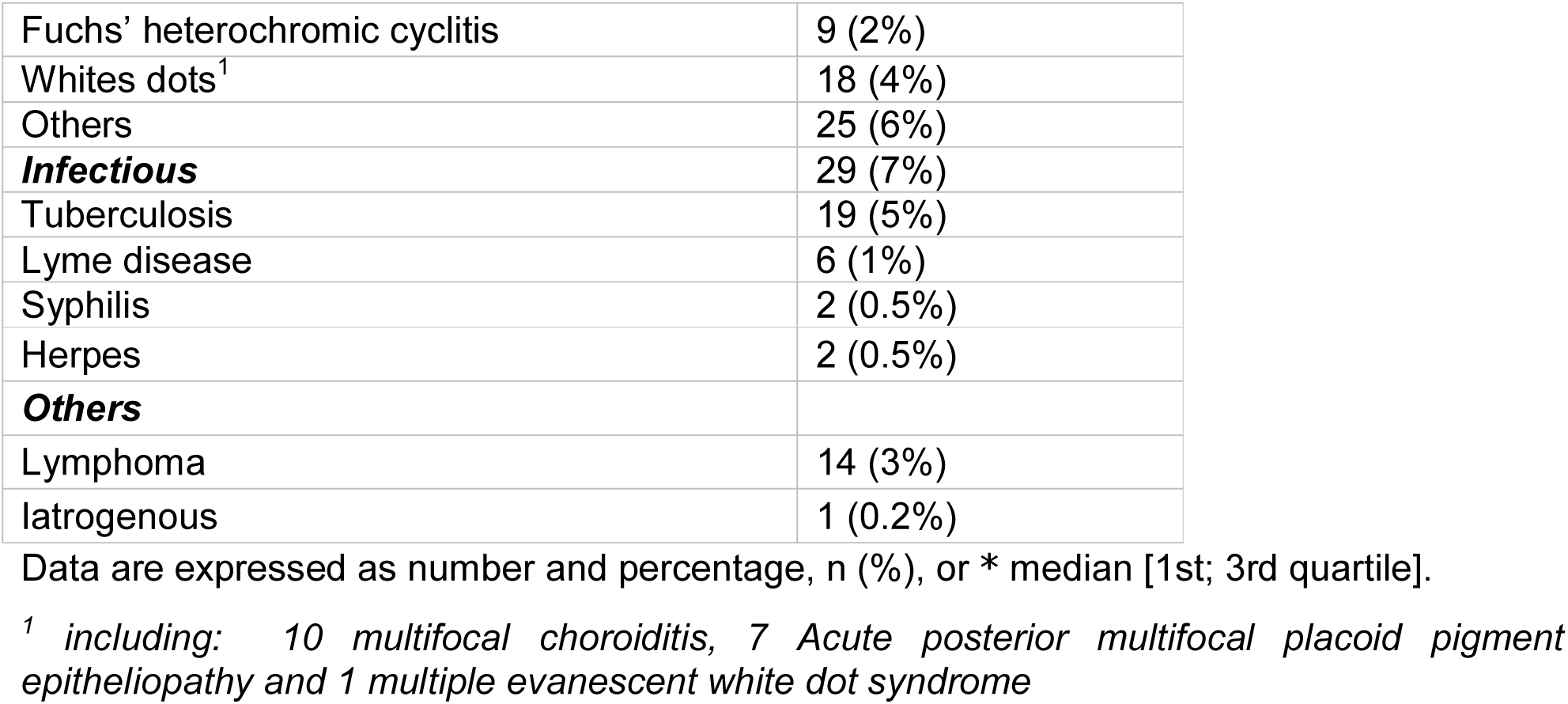
General characteristics, ophthalmological findings and etiological distribution in 402 patients

The anatomical distribution of uveitis was: anterior uveitis (AU, 11%), intermediate uveitis (IU, 30%), posterior uveitis (PU, 33%), or panuveitis (UP, 27%). Among the 402 patients, 68% had a bilateral disease and 84% had a chronic course. Most were non-hypertensive (94%) or non-granulomatous (77%). Unilateral papillitis was present in 83 cases (21%), and bilateral papillitis in 36 cases (9%). Thirty four patients (8%) had one or more neurological manifestations before or at the time of the bMRI test: 7 cranial nerve impairments, 7 motor impairments, 5 sensitive impairments, 4 optic neuropathies, 3 cerebellar syndromes, 2 states of mental confusions, 2 seizures, 1 frontal syndrome, 1 nystagmus, 1 peripheral neuropathy and 1 atypical headache.

### Relevance of bMRI

bMRI was considered to be abnormal in 80 cases (20%), including 26 with non-specific hypersignals (32%), 34 with vascular leucopathy (43%), 16 with demyelinating lesions (20%), and 4 with parenchymental mass (5%). bMRI was considered to be contributive in 19 cases (5%). An abnormal bMRI led to the diagnosing of 10 cases of MS, 5 RIS, and 4 oculocerebral lymphomas (OCL) (Table 2).

**Table 2:** General characteristics, ophthalmological findings of patients with a contributive cerebral

| Age | Sex | Uveitis anatomy | Ophthalmologic signs | B | C | G | Diagnosis | Neurological sign | bMRI | Treatment |
| --- | --- | --- | --- | --- | --- | --- | --- | --- | --- | --- |
| 20 | M | Pan-U | retinal vasculitis | X | X |  | MS | sensitive and motor impairment<br>optic neuropathy | inflammatory hypersignals<sup>1</sup> | yes (azathioprine) |
| 32 | M | IU |  | X | X | X | MS | motor impairment<br>optic neuropathy | inflammatory hypersignals<sup>1</sup> | yes (copaxone) |
| 26 | M | IU | retinal vasculitis | X | X |  | RIS |  | inflammatory hypersignals<sup>2</sup> | no |
| 36 | W | UP | retinal vasculitis | X | X |  | RIS |  | inflammatory hypersignals<sup>2</sup> | no |
| 67 | W | UP |  |  | X |  | OCL |  | parietal and occipital lymphoma | yes (chemotherapy + ocular radiotherapy) |
| 33 | M | IU |  | X | X | X | MS | motor impairment | inflammatory hypersignals<sup>1</sup> | yes (refused) |
| 35 | W | Pan-U | retinal vasculitis | X | X | X | MS | sensitive and motor impairment | inflammatory hypersignals<sup>1</sup> | yes (azathioprine) |
| 26 | W | IU | retinal vasculitis | X | X |  | MS | motor impairment | Inflammatory hypersignals<sup>1</sup> | yes (corticoïds boluses + azathioprine) |
| 57 | M | IU | retinal vasculitis |  | X |  | OCL | confusion | thalamic lymphoma | yes (chemotherapy) |
| 28 | W | IU | retinal vasculitis |  | X | X | RIS |  | inflammatory hypersignals<sup>2</sup> | no |
| 43 | W | Pan-U |  | X | X | X | RIS |  | inflammatory hypersignals<sup>2</sup> | no |
| 46 | W | Pan-U | retinal vasculitis<br>macular edema | X | X | X | MS | optic neuropathy | inflammatory hypersignals<sup>1</sup> | yes (azathioprine) |
| 60 | W | IU |  | X | X | X | OCL | frontal syndrome | frontal lymphoma | yes (chemotherapy) |
| 68 | W | IU |  | X | X | X | OCL | cerebellar syndrome | lymphoma | yes (unknown) |
| 38 | W | IU | periphlebitis |  | X |  | MS | sensitive impairment | inflammatory hypersignals<sup>1</sup> | yes (refused) |
| 29 | W | IU |  | X | X | X | MS | sensitive impairment | inflammatory hypersignals<sup>1</sup> | yes (mycophenolate mofetil) |
| 56 | W | Pan-U |  | X |  |  | RIS | 0 | inflammatory hypersignals<sup>2</sup> | no |
| 56 | W | IU | retinal vasculitis |  |  |  | MS | optic neuropathy | inflammatory hypersignals<sup>1</sup> | yes (azathioprine) |
| 43 | W | IU |  |  |  | X | MS | sensitive impairment | inflammatory hypersignals<sup>1</sup> | yes (azathioprine) |

A: acute, bMRI :brain magnetic resonance imaging; B: bilateral, G: granulomatous, MO: macular oedema ; MS: multiple sclerosis; OCL: oculocerebral lymphoma; RIS: radiologically isolated syndromes; RV: retinal vasculitis ; U: unilateral
<sup>1</sup>: According to 2010 revised McDonald's criteria
<sup>2</sup>: According to Okuda's criteria

Features associated with a contributive bMRI are summarized in Table 3. Among patients with a contributive bMRI, 13 (68%) had neurological signs, 9 (47%) had retinal vasculitis, and 1 (5%) had periphlebitis. In the non-contributive group, 21 patients (5%) had neurological signs, 61 (16%) presented retinal vasculitis, and one (0,3%) had periphlebitis. Cases with contributive MRI are detailed in Table 2.

**Table 3:**
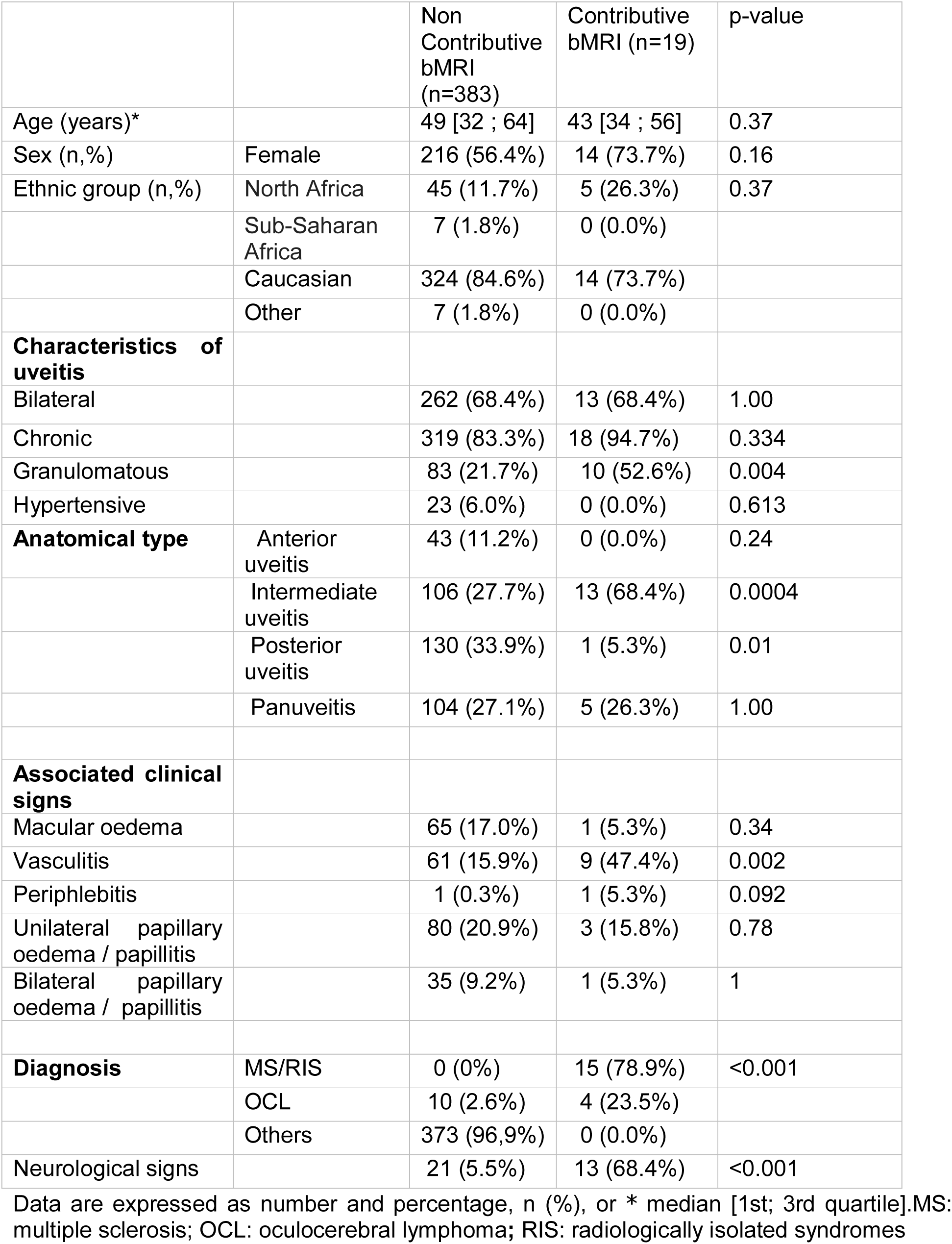
General characteristics and ophthalmological findings according to normal or abnormal bMRI (n=402)

|  |  | Non<br>Contributive<br>bMRI<br>(n=383) | Contributive<br>bMRI (n=19) | p-value |
| --- | --- | --- | --- | --- |
| Age (years)* |  | 49 [32 ; 64] | 43 [34 ; 56] | 0.37 |
| Sex (n,%) | Female | 216 (56.4%) | 14 (73.7%) | 0.16 |
| Ethnic group (n,%) | North Africa | 45 (11.7%) | 5 (26.3%) | 0.37 |
|  | Sub-Saharan<br>Africa | 7 (1.8%) | 0 (0.0%) |  |
|  | Caucasian | 324 (84.6%) | 14 (73.7%) |  |
|  | Other | 7 (1.8%) | 0 (0.0%) |  |
| **Characteristics of uveitis** |  |  |  |  |
| Bilateral |  | 262 (68.4%) | 13 (68.4%) | 1.00 |
| Chronic |  | 319 (83.3%) | 18 (94.7%) | 0.334 |
| Granulomatous |  | 83 (21.7%) | 10 (52.6%) | 0.004 |
| Hypertensive |  | 23 (6.0%) | 0 (0.0%) | 0.613 |
| **Anatomical type** | Anterior<br>uveitis | 43 (11.2%) | 0 (0.0%) | 0.24 |
|  | Intermediate<br>uveitis | 106 (27.7%) | 13 (68.4%) | 0.0004 |
|  | Posterior<br>uveitis | 130 (33.9%) | 1 (5.3%) | 0.01 |
|  | Panuveitis | 104 (27.1%) | 5 (26.3%) | 1.00 |
| **Associated clinical signs** |  |  |  |  |
| Macular oedema |  | 65 (17.0%) | 1 (5.3%) | 0.34 |
| Vasculitis |  | 61 (15.9%) | 9 (47.4%) | 0.002 |
| Periphlebitis |  | 1 (0.3%) | 1 (5.3%) | 0.092 |
| Unilateral papillary<br>oedema / papillitis |  | 80 (20.9%) | 3 (15.8%) | 0.78 |
| Bilateral papillary<br>oedema / papillitis |  | 35 (9.2%) | 1 (5.3%) | 1 |
| **Diagnosis** |  |  |  |  |
|  | MS/RIS | 0 (0%) | 15 (78.9%) | <0.001 |
|  | OCL | 10 (2.6%) | 4 (23.5%) |  |
|  | Others | 373 (96,9%) | 0 (0.0%) |  |
| Neurological signs |  | 21 (5.5%) | 13 (68.4%) | <0.001 |

Data are expressed as number and percentage, n (%), or \* median [1st; 3rd quartile].MS: multiple sclerosis; OCL: oculocerebral lymphoma; RIS: radiologically isolated syndromes

Uveitis remained of undetermined origin in 27 cases in spite of bMRI abnormalities. Among patients with neurological signs, 13 (38%) had a contributive bMRI. In the absence of neurological signs, only 6 patients had a contributive bMRI with 1% leading to a diagnosis: 5 RIS, and 1 OCL. Only the patient with OCL received a specific treatment after this diagnosis.

In the univariate analysis, a contributive bMRI was associated with the following features: intermediate uveitis (p<0.001), presence of retinal vasculitis (p=0.002), granulomatous type (p=0.003), and presence of neurological signs (p<0.001) (Table 4).

**Table 4:** Results of univariate and multivariate analyses

|  | Univariate analysis |  | Multivariate analysis |  |
| --- | --- | --- | --- | --- |
|  | OR (95% CI) | P-value | OR (95% CI) | p-value |
| Intermediate uveitis | 5.66 (2.10-15.3) | <0.001 | 4.33 (1.42-13.17) | 0.010 |
| Posterior uveitis | 0.11 (0.014-0.82) | 0.031 |  |  |
| Retinal vasculitis | 4.75 (1.85-12.20) | 0.002 |  |  |
| Periphlebitis | 21.2 (1.44-401.3) | 0.033 |  |  |
| Granulomatous | 4.02 (1.58-10.2) | 0.003 |  |  |
| Neurological signs | 37.3 (12.9-108.1) | <0.0001 | 32.01 (10.71-95.64) | <0.001 |

In the multivariate regression analysis, the main features independently associated with having a contributive bMRI were: intermediate uveitis (p=0.01) and the presence of neurological signs (p<0.001) (Table4).

Patients with MS and RIS-related uveitis were non-significantly younger (p=0.05), with a median age of 38. While patients with OCL were significantly older (p<0.001) with a median age of 67, they were all above 40 years of age (Table 5), ranging from 55 to 88 years old.

**Table 5:** Oculocerebral lymphoma and multiple sclerosis characteristics compared to general population

|  | **Total** | **OCL** |  | **MS / RIS** |  |
| --- | --- | --- | --- | --- | --- |
|  | N = 402 | N = 14 | **p-value** | N = 15 | **p-value** |
| Age (years) * | 49 [33 ; 63] | 67 [60 ; 74] | <0.001 | 38 [34 ; 44] | 0.050 |
| Sex (F) | 230 (57.2%) | 8 (57.1%) | 1 | 11 (73.3%) | 0.29 |
| **Ethnicity** |  |  |  |  |  |
| North Africa | 50 (12.4%) | 3 (21.4%) | 0.64 | 5 (33.3%) | 0.18 |
| Sub-Saharan Africa | 7 (1.7%) | 0 (0.0%) |  | 0 (0.0%) |  |
| Caucasian | 338 (84.1%) | 11 (78.6%) |  | 10 (66.7%) |  |
| Other | 7 (1.7%) | 0 (0.0%) |  | 0 (0.0%) |  |
| **Anatomical type** |  |  |  |  |  |
| UA | 43 (10.7%) | 0 (0.0%) | 0.38 | 0 (0.0%) | 0.39 |
| UI | 119 (29.6%) | 7 (50.0%) | 0.13 | 10 (66.7%) | 0.003 |
| UP | 131 (32.6%) | 2 (14.3%) | 0.16 | 0 (0.0%) | 0.004 |
| PU | 109 (27.1%) | 5 (35.7%) | 0.54 | 5 (33.3%) | 0.56 |
| **Characteristics of uveitis** |  |  |  |  |  |
| Bilateral | 275 (68.4%) | 11 (78.6%) | 0.56 | 11 (73.3%) | 0.78 |
| Chronic | 336 (83.5%) | 13 (92.9%) | 0.71 | 14 (93.3%) | 0.48 |
| Granulomatous | 93 (23.1%) | 1 (7.1%) | 0.2 | 10 (66.7%) | < 0.001 |
| Hypertensive | 23 (5.7%) | 1 (7.1%) | 0.57 | 0 (0.0%) | 1 |
| **Associated clinical signs** |  |  |  |  |  |
| Macular oedema | 66 (16.4%) | 3 (21.4%) | 0.71 | 1 (6.7%) | 0.48 |
| Retinal vasculitis | 70 (17.4%) | 2 (14.3%) | 1 | 8 (53.3%) | 0.001 |
| Periphlebitis | 2 (0.5%) | 0 (0.0%) | 1 | 1 (6.7%) | 0.073 |
| Unilateral papillary oedema / papillitis | 83 (20.6%) | 2 (14.3%) | 0.74 | 3 (20.0%) | 1 |
| Bilateral papillary oedema / papillitis | 36 (9.0%) | 0 (0.0%) | 0.63 | 1 (6.7%) | 1 |
| Contributive bMRI | 19 (4.7%) | 4 (28.6%) | 0.003 | 15 (100%) | <0.001 |
| Neurological signs | 34 (8.5%) | 4 (28.6%) | 0.023 | 10 (76.7%) | <0.001 |

Data are expressed as number and percentage, n (%), or \* median [1st; 3rd quartile]. AU: anterior uveitis; bMRI: brain magnetic resonance imaging; IQ: Interquartile; IU: intermediate uveitis; MS: multiple sclerosis; OCL: oculocerebral lymphoma; PU: posterior uveitis; RIS: radiologically isolated syndromes; UP: panuveitis

## Discussion

The value of bMRI for the etiological diagnosis of uveitis is not well defined. In the present study, we found a minority (5%) of patients who underwent a bMRI had abnormalities contributive to etiological diagnosing. Among subjects free of neurological signs, bMRI was abnormal in only few cases (1%), leading to a diagnosis of either RIS or OCL. Since bMRI is an expensive exam [8] sometimes associated with side effects such as allergy and not always easily accessible, it should not be prescribed for all patients with unexplained uveitis. In a previous study, Hadjadj et al. reported that the presence of snowballs in the vitreous humor and retinal vasculitis were among the features associated with abnormal bMRI [9]. Because abnormal MRI features are often nonspecific, we chose to assess the relevance of MRI abnormalities in the etiological diagnosing of uveitis. Our work shows that intermediate uveitis, retinal vasculitis, granulomatous uveitis, and the presence of neurological signs were all significantly related to a contributive bMRI.

In the ULISSE study [11], a bMRI was performed in 44 patients, and was contributive in 16% of the cases. A retrospective study of 71 patients with intermediate/posterior uveitis, or panuveitis showed that a systematic bMRI was contributive in 8% of the cases [14]. Hadjadj et al. reported similar results: a brain MRI was performed in 168 patients and was contributive in 15 patients (8.9%), among whom only three presented neurological symptoms. Abnormal findings were most often demyelinating lesions. They reported that bMRI led to the diagnosis of neurosarcoidosis in 4 cases and Behçet’s disease in 1 patient. Like other authors, we consider that asymptomatic demyelinating lesions in patients with sarcoidosis and Behçet’s disease should not lead to the start of a systemic treatment.

In our study, MS was the main diagnosis in patients with contributive bMRI. For the most part, uveitis is known to precede the diagnosis of MS [26]. However only 1% of MS uveitis patients will present an uveitis, which explains why it is not used as a diagnostic criterion for MS. Several studies have shown a prevalence of MS ranging from 7% to 30% in patients with intermediate uveitis, leading the authors to suggest systematic bMRI for these patients [27].

For Petrushkin et al. bMRI should not be proposed in the absence of a history or clinical signs suggestive of MS, nor in the absence of prognostic or therapeutic consequences [27]. Five of our contributive bMRI tests led to the diagnosing of RIS. Among this subgroup of patients who have asymptomatic demyelinating lesions which are highly suggestive of MS, one-third may convert to clinically-established MS within 5 years [28]. RIS diagnosis today leads to a strict follow-up alone, although some trials (TERIS and ARISE studies) are ongoing to evaluate MS oral therapy in these cases so as to delay the onset of the first clinical event [29,30].

OCL is a rare disease that can manifest as an intermediate/posterior uveitis or as panuveitis. It is thus essential to rule out this diagnosis in potential cases because OCL is a life-threatening condition. In suspected cases, a bMRI is generally suggested (with T1 sequences before and after injection of contrast material) to explore a lymphomatous parenchymal mass. In our study, 4 patients had OCL with a contributive MRI, including 3 patients with neurological symptoms. All patients were over 40 and had chronic intermediate/posterior uveitis or panuveitis.

In a previous study, we had developed recommendations for the diagnosing of uveitis based on a review of the literature and the findings of the ULISSE study [31]. We proposed that bMRI should be reserved for patients over 40 presenting unexplained uveitis, when searching for cerebral lesions suggestive of OCL. In our study OCL patients were significantly older than the general population, and this therefore reinforces our previous recommendations.

In addition to these recommendations, Winkelmann al. recommended performing bMRI before testing for monoclonal anti-TNFα antibodies, due to the risk of aggravating a demyelinating disease [32]. For the same reasons, Wakefield et al. recommended that bMRI be done before giving anti-TNFα or anti-IL6 therapy to patients with intermediate uveitis [33]. In our study, bMRI frequently pinpointed hypersignals which were either non-specific, or linked to white matter hyperintensities of vascular origin or inflammatory lesions. Nonetheless bMRI interpretation in these cases can be difficult and may depend on the operator, with frequent errors at risk. Neurological expertise and lumbar puncture should be offered, to examine the link between MRI abnormalities and uveitis etiology. Nearly 99% of the bMRIs performed in the absence of neurological signs were non-contributive. Patients with no neurological signs but with contributive bMRIs can be split into two groups: the first covers RIS patients with no specific treatment requirements, and the second covers OCL patients who were all older than 40.

Our work established an inverse correlation between posterior uveitis and a contributive MRI. This may be explained by a small number contributive MRIs, since MS/RIS and OCL uveitis are usually intermediate [34,35]. Moreover, as recommended by the SUN [15], uveitis was classified as intermediate when the vitreous was the major site of inflammation even if there was posterior uveitis. However, since posterior and panuveitis are common manifestations of OCL, it appears essential in these situations to rule out this etiology.

There are some limitations to our study. First, this study is retrospective and monocentric, and bMRI was not performed in all cases. Second, because of its retrospective nature this study suffers from a reporting bias: indeed the clinical examination was not standardized, and this may explain the low rate of papillary edema, cystoid macular edema and periphlebitis. Third, since our center is a tertiary referral center for the management of uveitis, some patients had a long history of uveitis before being referred to us, and thus could differ from daily-care patients. However, this last point also made it possible to include patients with sufficient follow-up after initial diagnosis. Fourth, patients in this study were referred by ophthalmologists to internists according to no pre-established criteria, but mainly for cases of ophthalmological presentation suggestive of systemic disease. This may have biased the patient selection, leading to over-representation of systemic diseases. Finally, the bMRI analysis was not standardised by a radiologist blinded to the patient’s clinical history.

## Conclusion

In conclusion, our study shows that in the context of etiological uveitis diagnosing, bMRI is relevant in two specific situations: in patients with neurological signs and/or retinal vasculitis, and in patients older than 40 years with intermediate/posterior uveitis or panuveitis. Given the cost of bMRI, we suggest limiting examinations to patients presenting with at least one of these features. The most important diagnoses to look for are multiple sclerosis, in the first case, and oculocerebral lymphoma in the second. Further controlled studies with larger numbers of patients are needed to confirm our results and to clarify the clinical usefulness of bMRI in patients with uveitis of unknown origin.

## Data Availability

These are anonymised patient data, owned by Nicolas Chirpaz and reuse is not permitted.

## Contributors

NC, PS, TM and AG were the principal investigators who conceived and designed the study. NC and RB collected data. SK performed statistical analysis. All authors interpreted the data and approved the manuscript. NC and AG drafted the manuscript. PS, TM, YJ, SK and LK revised the manuscript.

## Ethics approval

This study was approved by the ethics committee of the French Society of Ophtalmology (IRB 00008855 Société Française d’Ophtalmologie IRB#1)

